# Biological, Behavioral, Environmental and Clinical Characteristics of Vitiligo Patients at Gampaha Wickramarachchi Ayurveda Teaching Hospital, Yakkala, Sri Lanka- Descriptive cross-sectional study

**DOI:** 10.1101/2025.08.24.25334308

**Authors:** Fathima Shazmin Hazari

**Author notes:** **Corresponding author at:** Faculty of Graduate Studies, University of Kelaniya, Sri Lanka.

## Abstract

**Background:** Vitiligo is a complex skin disorder with multifactorial etiology, including genetic, environmental, and behavioral factors. In Sri Lanka, data on the epidemiology and characteristics of Vitiligo patients, particularly in Ayurvedic clinical settings, remain limited.

**Objectives:** This study aimed to explore the biological, behavioral, environmental, and clinical characteristics of vitiligo patients receiving Ayurvedic treatment at the Gampaha Wickramarachchi Ayurveda Teaching Hospital, Sri Lanka, and to examine associations with comorbid conditions.

**Methods:** A total of 108 patients with clinically diagnosed Vitiligo were consecutively recruited. Data on socio-demographic, behavioral, environmental, and clinical variables were collected using a structured interviewer-administered questionnaire. Descriptive statistics and correlation analyses were performed to identify key patterns and associations.

**Results:** The majority of patients were female (60.9%) with a mean age of 44 years, and 17.3% reported a family history of Vitiligo. Most resided in the Western Province, exhibited dietary imbalances, and reported behavioral factors such as low exercise and betel chewing. Localized vitiligo types predominated, with common lesions on upper and lower limbs. Comorbid dermatological allergies and systemic conditions were frequent, with hyperlipidemia significantly correlated with Vitiligo type (p = 0.041). Other comorbidities showed no significant association.

**Conclusions:** This study provides important local epidemiological data, revealing a multifactorial profile of Vitiligo patients in an Ayurvedic setting. The significant link between hyperlipidemia and Vitiligo suggests potential metabolic involvement. Integration of traditional and biomedical approaches may enhance patient management. Larger, multi-center studies are needed to validate and extend these findings.

- This study used a structured interviewer-administered questionnaire, enhancing data completeness and consistency.
- Consecutive recruitment of participants minimized selection bias within the clinic population.
- Data collection included a comprehensive range of biological, behavioral, environmental, and clinical variables for a holistic understanding.
- The single-center, cross-sectional design limits causal inference and may affect the generalizability of findings to broader populations.
- Self-reported behavioral and environmental data may be subject to recall bias, which could impact accuracy.

## Introduction

Vitiligo is a chronic, acquired skin disorder characterized by the progressive loss of skin pigmentation due to the destruction of melanocytes^1^. This is actuated through complicated and multifactorial pathways that include genetic susceptibility, autoimmune responses, oxidative stress and environmental triggers^2^. Clinically, the condition manifests in well-defined areas of white patches or lesions on the body; thus, it may be accompanied by itching and burning sensation^3^. The condition can develop in young age, with a substantial number of cases initiated at age 20 or below^4^. Multiple risk factors including genetic predisposition, immune dysfunction, sun exposure, chemical or physical trauma, and lifestyle influences contribute to its onset and progression of Vitiligo, although the precise causes often remain unclear ^5,6,7^

The fact that limited improvement occurs in response to therapy and the time consumed in such re-pigmentation further exacerbate the psychological impact of the disease, often forcing patients to conceal their Vitiligo lesions to improve their confidence and self-esteem ^8,9^. Such psychosocial burdens can significantly affect the daily functioning, interpersonal relationships, and treatment-seeking behavior in affected individuals ^10,11^Although extensive research on the biological, behavioral, and environmental factors of Vitiligo abounds, most of the findings are context-specific and centered on populations outside South Asia ^12,13^. This constrains the external validity of such studies to various contexts such as Sri Lanka ^14.^ Moreover, there is a notable absence of research focusing on the epidemiological and clinical characteristics of Vitiligo patients within the Sri Lankan healthcare context, particularly in an Ayurvedic medical setting.

The Gampaha Wickramarachchi Ayurveda Teaching Hospital in Yakkala, Sri Lanka, is one of the foremost educational and clinical institutions of traditional medicine under the University of Indigenous Medicine ^15.^ It has combined both Ayurvedic and conventional ways of treating Vitiligo, providing it with a unique environment to explore Vitiligo in both biomedical and indigenous concepts. This descriptive cross-sectional study is appropriate for documenting the prevalence, characteristic demographic distribution, and clinical manifestations patients of Vitiligo at Gampaha Wickramarachchi Ayurveda Teaching Hospital. This study aimed to describe the biological, behavioral, environmental, and clinical characteristics, of patients with Vitiligo. It also attempted to investigate the relationships between comorbid conditions, and Vitiligo patch distribution. These features are more comprehensible to allow the designing of patient-centered managerial approaches and preventive measures. Furthermore, early identification of potential complications and comorbidities may improve clinical outcomes and quality of life for Vitiligo patients.

## Methods

### Study Design and Setting

The research was a descriptive cross-sectional study that lasted (April-July 2025) at the Skin Clinic of Gampaha Wickramarachchi Ayurveda Teaching Hospital, in Yakkala, Sri Lanka. Key outcomes and exposures, predictors, possible confounders, and impact modifiers had been defined before data were collected. Vitiligo was clinically diagnosed according to the standard diagnostic criteria through a clinical examination concurred by the researcher in collaboration with the in-charge consultant at the Skin Clinic.

Certain behavioral and environmental factors which include: diet, residence, working conditions, educational level, smoking and exposure to sun were measured using a structured questionnaire which the researcher conducted. Comorbidity was checked by examining medical records where possible. All participants were interviewed using the same standardized questionnaire, ensuring consistency in measurement methods

To reduce potential selection bias, all eligible patients attending the clinic during the study period who met the inclusion criteria were consecutively invited to participate. Interviewer administered questionnaires were useful in minimizing information bias as there was the capacity to clarify with the participants. Recall bias was recognized as a limitation due to reliance on self-reported behavioral data.

### Study Population and Sampling

Clinically diagnosed patients with Vitiligo, aged between 15 to 80 years, both sexes, and patients with second attempt treatment and were started on Ayurvedic medicine were recruited according to predefined inclusion criteria. Pregnant, lactating women and individuals unwilling to participate were excluded. The required sample size (n = 108) was computed based on a 95% confidence level, 5% margin of error, and adjustment for the finite population.

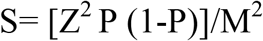

Quantitative data represented by age and the duration of the disease were treated both as continuous variables and as categories defined by clinical relevant (age groups: 15-24, 25-34,35-44, >44 years). Groupings were chosen to facilitate meaningful comparisons and align with similar Vitiligo studies.

### Data Collection

Data were collected using a structured, interviewer-administered questionnaire including three main sections: (1) socio-demographic information, which included biological (e.g., age, gender, religion), environmental (e.g., residence, marital status, employment, family system, exposure to environmental triggers), and behavioral determinants (e.g., psychiatric history, diet, bowel movements, sleep patterns, habits related to the disorder); (2) disease-related characteristics, encompassing comorbidities and details of lesions; and (3) clinical determinants related to vitiligo and associated health conditions.

### Ethical Considerations

Ethical approval was obtained from the Ethics Review Committee, Faculty of Medicine, University of Kelaniya, Ragama, Sri lanka. (Ref: P/127/09/2024). Written informed consent was obtained from all participants after explaining the study purpose, procedures, and voluntary nature of participation.

### Data Analysis

Statistical analysis was done through SPSS with descriptive statistics, independent-sample t-test, and one way ANOVA, as appropriate. Statistical test assumptions were verified and p<0.05 was adopted as a significance level. Possible confounding factors were discussed through the calculation of all the percentages of biological, behavioral, environmental and clinical determinants. Correlation analysis was done to target specifically the extent to which the comorbid diseases are associated with varied Vitiligo. There were very few missing data which were filled using list wise deletion. Sensitivity analysis was not performed because the study was of descriptive nature and because of completeness of the data.

### Patient and Public Involvement

Patients were involved in the design of the research through consultation meetings. Their feedback helped shape the research questions and patient information materials. They were engaged throughout the study to advice on recruitment strategies and dissemination plans.

## Results

There were 120 patients who were screened at the beginning of the study period. Among them, 12 were excluded. Thus, 108 patients with clinically diagnosed Vitiligo were included. The non-participation was due to reasons such as e.g., refusal, pregnancy, and incomplete data.

### Biological characteristics

The majority were female (60.9%), with the average age of 44 years, Most participants were Buddhist (82%) and 8.7 percent had documented history of psychiatry (Table 1).

**Table 1.** Biological characteristic features of individuals with Vitiligo.

| Determinant | Category | n | % |
| --- | --- | --- | --- |
| Age (years) | 15–24 | 14 | 13 |
|  | 25–34 | 9 | 8 |
|  | 35–44 | 33 | 31 |
|  | >44 | 52 | 48 |
| Gender | Male | 42 | 39.1 |
|  | Female | 66 | 60.9 |
| Religion | Buddhism | 89 | 82 |
|  | Catholic | 5 | 5 |
|  | Islam | 14 | 13 |
| Psychiatric history | No | 99 | 91.3 |
|  | Yes | 9 | 8.7 |

### Environmental and Behavioral Characteristics

Most participants resided in the Western Province (82.6%), were married (78%), and lived in nuclear families (56%). Over half (52%) had completed secondary education. Occupationally, the majority were housewives (17.4%) and office workers (13%). Environmental triggers included animal bites (22.7%), rat urine contamination (4.3%), cosmetic/medication allergies (25.9%), chemical exposures (4.3%), and sunlight (8.7%) (Table 2). Behavioral patterns included imbalanced diets (69.5%), low water intake (78.3%), Of these, 60.9% had mostly healthy bowel motions and mostly normal sleep (73.9%). Exercise (17.4%) and chewing betel (17.4%) were listed under the patients’ behaviors (Table 3).

**Table 2.** Environmental characteristic features of individuals with Vitiligo.

| **Determinant** | **Category** | **n** | **%** |
| --- | --- | --- | --- |
| **Residence** | Western Province | 89 | 82.6 |
|  | Northwestern Province | 5 | 4.4 |
|  | Southern Province | 14 | 13 |
| **Marital status** | Single | 19 | 17.5 |
|  | Married | 84 | 78 |
|  | Separate | 5 | 4.5 |
| **Education** | Primary | 23 | 21 |
|  | Elementary | 20 | 19 |
|  | Secondary | 56 | 52 |
|  | Tertiary | 9 | 8 |
| **Employment** | Housewife | 19 | 17.4 |
|  | Office workers | 14 | 13 |
|  | Teachers/Health workers | 9 | 8.7 |
|  | Shopkeepers/Labor/Farmers/Business/Drivers etc. | 39 | 36.1 |
|  | Students | 3 | 2.8 |
| **Family system** | Joint | 42 | 39 |
|  | Nuclear | 61 | 56 |
|  | Alone | 5 | 5 |
| **Trigger factors** | Rat bite / AC exposure | 14 | 13 |
|  | Others (Gloriosa juice, Sun exposure, Dog bite, Medicines, etc.) | 9<br>5 | 8.7<br>4.3 |

**Table 3.** Behavioral characteristic features of individuals with Vitiligo.

| **Determinant** | **Category** | **n** | **%** |
| --- | --- | --- | --- |
| **Diet habits** | Irregular | 9 | 8.7 |
|  | Imbalanced | 75 | 69.6 |
|  | Balanced | 24 | 21.7 |
| **Water intake** | Good | 23 | 21.7 |
|  | Poor | 85 | 78.3 |
| **Bowel movement** | Normal | 66 | 60.9 |
|  | Constipated | 37 | 34.8 |
|  | Irregular | 5 | 4.3 |
| **Sleeping pattern** | Normal | 80 | 73.9 |
|  | Disturbing | 28 | 26.1 |
| **Habits** | Betel chewing | 19 | 17.4 |
|  | Exercise | 9 | 8.7 |
|  | Smoking | 5 | 4.3 |
|  | None | 75 | 69.6 |

**Table 4.** Clinical characteristic of individuals with Vitiligo.

| **Clinical characteristic** | **Category** | **n** | **%** |
| --- | --- | --- | --- |
| **Type of Vitiligo** | Local | 42 | 39.1 |
|  | Generalized | 28 | 26.1 |
|  | Segmental | 19 | 17.4 |
|  | Acro-facial | 14 | 13 |
|  | Mucosal | 5 | 4.3 |
| **Sites involved** | Face | 11 | 10 |
|  | Fingers | 37 | 34.8 |
|  | Lips | 47 | 43.4 |
|  | Scalp | 14 | 13 |
|  | Upper limb | 70 | 65.1 |
|  | Lower limb | 56 | 52.1 |
|  | Trunk | 23 | 21.7 |
|  | Full body | 9 | 8.7 |
| **Duration of disease** | 1–6 months | 14 | 13 |
|  | 6–12 months | 19 | 17.4 |
|  | 1–3 years | 33 | 30.4 |
|  | >3 years | 42 | 39.1 |
| **Symptoms** | Itching | 19 | 17.4 |
|  | No itching | 89 | 82.6 |
| **Family history** | Within family | 14 | 13 |
|  | Relatives | 5 | 4.3 |
|  | None | 89 | 82.6 |
| **Allergic conditions** | Mosquito bite allergy | 30 | 27.8 |
|  | Imitation jewelry allergy | 21 | 19.4 |
|  | Food allergy | 9 | 8.3 |
|  | Others | 4 | 3.7 |
|  |  | 5 | 4.6 |
| **Previous treatment** | Ayurveda | 14 | 13 |
|  | Western | 47 | 43.5 |
|  | Mixed | 5 | 4.6 |
|  | No treatment | 42 | 39.1 |
| **Other diseases** | Rheumatoid arthritis | 23 | 21.7 |
|  | Hyperlipidemia | 9 | 8.6 |
|  | Wheezing | 9 | 8.6 |
|  | Hypertension | 9 | 8.6 |
|  | Hypothyroidism | 5 | 4.3 |

**Table 5.** Significant Correlation between other disease and types of Vitiligo patches.

| Other illness | P value |
| --- | --- |
| Rheumatoid arthritis | 0.306 |
| Hyper-lipidemia | 0.041 |
| wheezing | 0.784 |
| Hypertension | 0.246 |

### Clinical Characteristics

According to the results, 39.1% of the patients had been diagnosed with a condition that had lasted more than three years. The body types with the highest and lowest percentages of Vitiligo involvement were mucosal (4.3%) and local types (39.1%) (figure1). Upper limb (65.1%), lower limb (52.1), lips (43.4%) were the body parts most involved, while the entire body and face were the least involved and gradually onset. Wounds accounted for 17.4% before appear white patches, followed by itching. Family history (17.3%) within the family (13%) and 4.3% were related generation. The majority of them had other dermatological conditions such as mosquito bite allergy (30%), imitation jewelry allergy (21.7%), and food allergy (8.6%), and they were medicated for rheumatoid arthritis (21.7%), hyperlipidemia, wheezing, hypertension, wheezing (25.8%), and thyroxin for hypothyroidism (4.3%). Most (43.5%) had previously received western therapy before attending the clinic.

**Figure 1.**
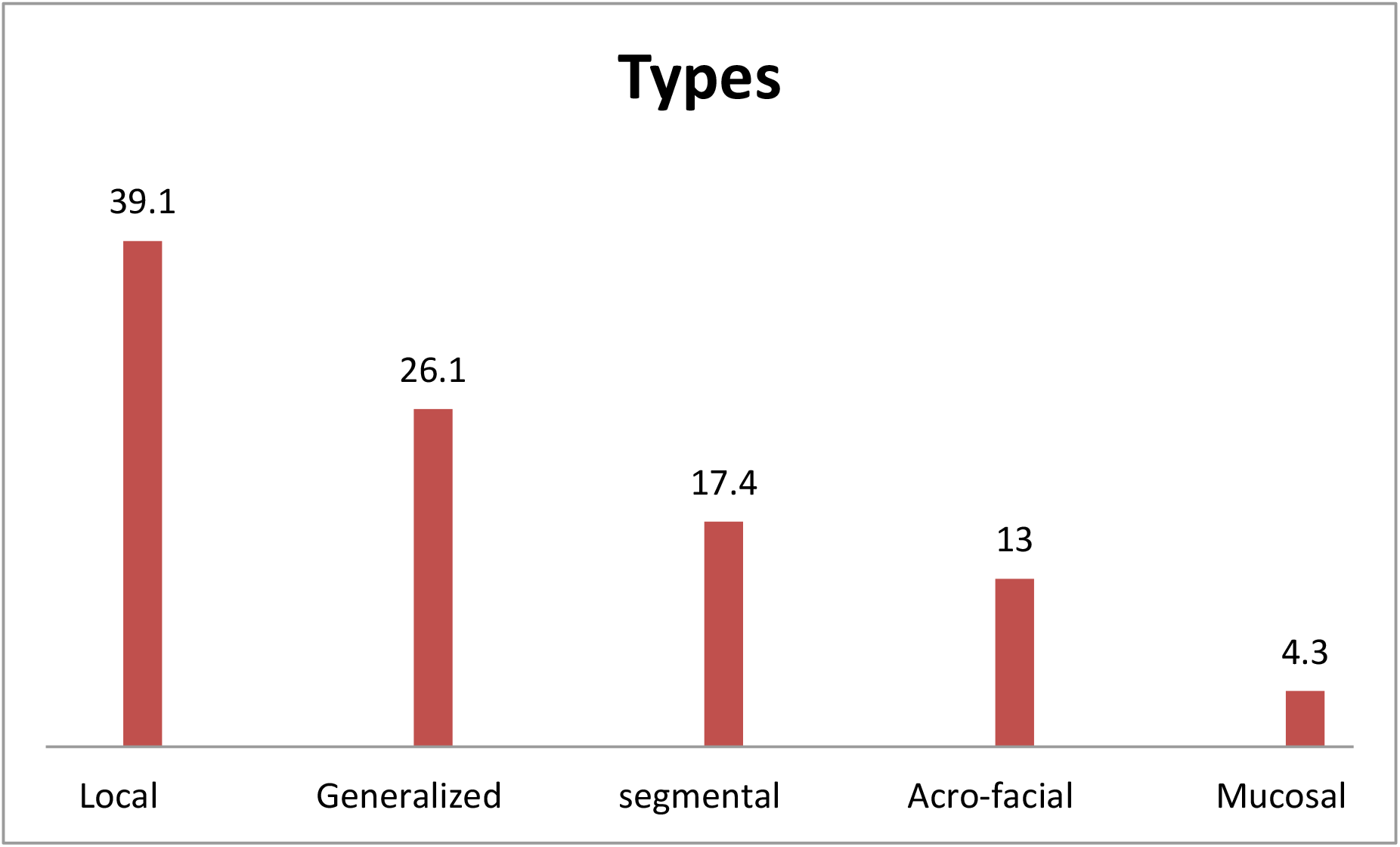
Types of Vitiligo

**Figure 2.**
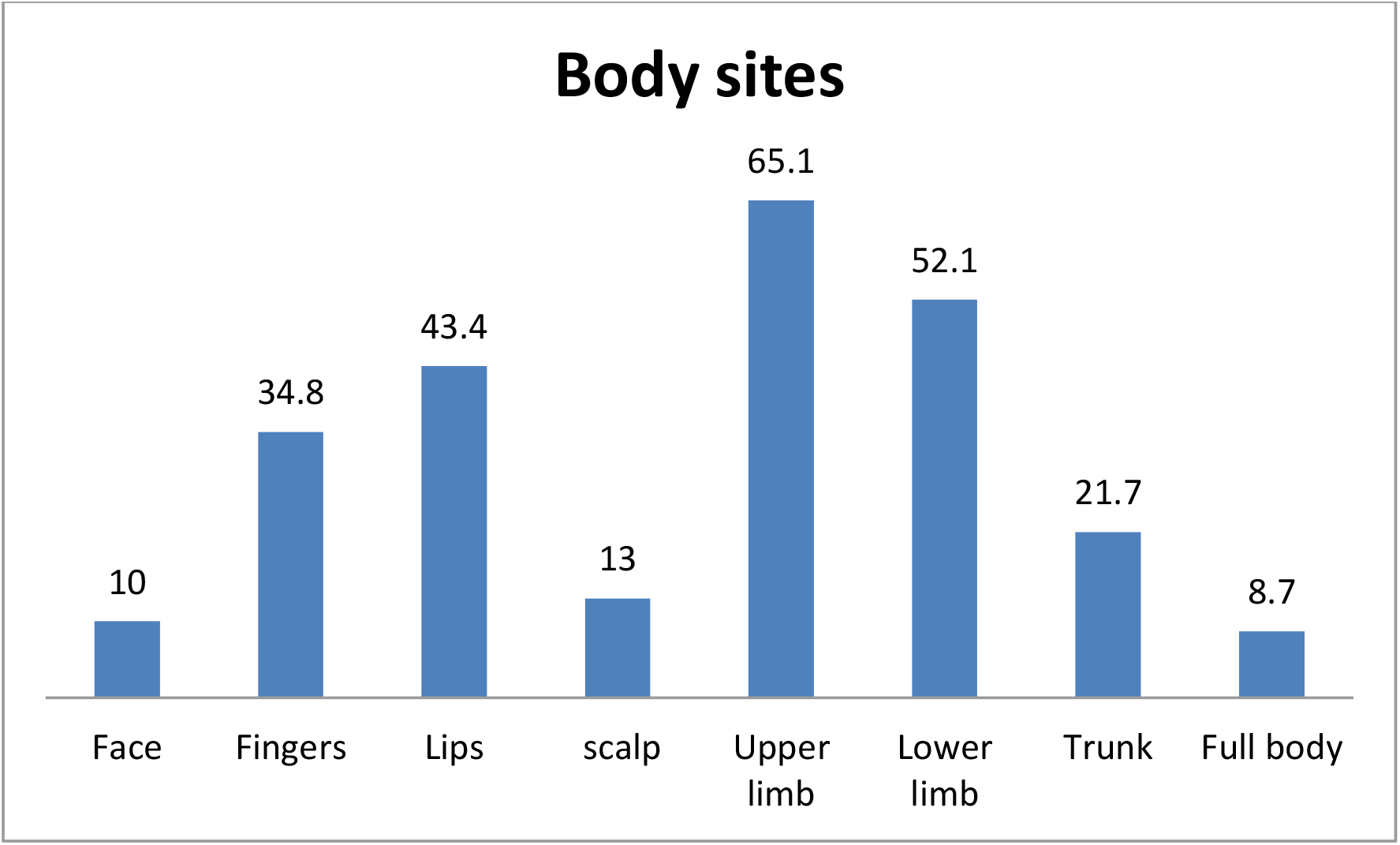
body sites

### Missing Data

Missing data were minimal; for all key variables, the number of participants with missing information was less than 5%. Variables with missing data are indicated in the corresponding tables.

### Associations with comorbid

The analysis of correlations between selected clinical factors, in patients with Vitiligo, regarding comorbid conditions, only hyperlipidemia showed a significant correlation with Vitiligo type (p = 0.041), whereas rheumatoid arthritis (p = 0.306), wheezing (p = 0.784), and hypertension (p = 0.246) were not significantly associated. Confidence intervals were not calculated due to the descriptive nature of the analysis.

## Discussion

This study explored the characteristics into biological, behavioral, environmental, and clinical aspects of 108 Vitiligo patients that visited the n Ayurvedic skin clinic in Sri Lanka, which would complement the existing system in the country that gap in local epidemiological data^16,17^. Consistent with prior research, females comprised the majority, and nearly half were over 44 years old. The prevalence of family history and psychiatric conditions are consistent with global evidence emphasizing genetic and psychosocial influences on Vitiligo^18,19,20,21^. Environmental factors such as residence primarily in the Western Province reflect the hospital’s location, while occupational, behavioral, and Ayurvedic concepts provide additional context for disease triggers^22,23,24^.

Behavioral findings revealed prevalent dietary imbalances, poor hydration, and lifestyle habits with low exercise and betel chewing which has been identified to exacerbate oxidative stress and immune suppression associated with Vitiligo progression ^25,26,27,28^. Clinical presentations varied in localized to mucosal types with common lesion sites consistent with literature, though some site patterns differed^29,30.^ Associations with comorbid dermatological allergies and systemic conditions were noted, with hyperlipidemia demonstrating significant correlation with Vitiligo type and emphasizing an apparent metabolic factor in the pathogenesis of the disease^31,32,33,34^.

The single center study design also forms a limitation to this study as it may constitute a challenge to generalizability to other populations outside the clinic. Although the factors of selection bias were criticized as limited by consecutive recruitment, the factors of the bias of recall potentially existed due to self-reported behavioral data. The descriptive cross-sectional nature precludes causal inference, and missing data were minimal but not zero, handled via list wise deletion. Multiplicity of analyses was addressed through conducting the analysis on defined variables but sensitivity analyses were not applied

Interpretation of findings should consider these limitations alongside existing evidence. The predominance of the female population and family history is suggestive of genetics and psychosocial factors towards the cause of Vitiligo in this population. Behavioral and environmental factors underscore the multifactorial nature of the disease and the potential role of Ayurveda-specific contributors. The significant correlation between hyperlipidemia and Vitiligo types justifies for integrating metabolic monitoring in patient management and further research on lipid metabolism’s role in Vitiligo.

The results have relevant implications for public health and clinical practice in Sri Lanka, specifically in integrating traditional Ayurvedic perspectives with biomedical approaches. While the single-center setting limits broad extrapolation, findings contribute valuable local epidemiological data.. In further studies, multi-site longitudinal design with larger samples enhances generalizability and explores causal pathways, including metabolic and psychosocial mechanisms influencing vitiligo. Additionally, public awareness campaigns are essential to dispel myths and superstitions surrounding vitiligo

## Conclusion

This study provides valuable insights into the biological, behavioral, environmental, and clinical characteristics of the patients with Vitiligo in an Ayurvedic clinical setting in Sri Lanka. The results highlight a predominance of female patients, significant genetic as well as psychosocial factors, and lifestyle that may affect the progression of the disease. The observed association between hyperlipidemia and Vitiligo type which indicates the presence of a possible metabolic aspect warranting further investigation. Despite limitations related to the single-center design and cross-sectional nature, the results emphasize the importance of integrating traditional and biomedical approaches for holistic patient management. Future research with larger, multi-site cohorts is needed to confirm these findings and explore causal mechanisms.

## Data Availability

The data that support the findings of this study are from the corresponding author upon reasonable request

## Author contributions

F.S Hazari-Conceptualization, Validation, Formal analysis, Writing – Original Draft.

## Acknowledgements

I would like to express my thanks to the director of the Gampaha Wickramarachchi Ayurveda Teaching Hospital in Yakkala, Sri Lanka, for her care in allowing me to conduct my study in the skin clinic.

## Patient consent for publication

Not applicable

## Data availability

### Declaration of Generative AI and AI-assisted technologies in the writing processes

During the preparation of this work the authors used Perplexity in order to assist with English writing and language refinement. After using this tool, the authors reviewed and edited the content as needed and take full responsibility for the content of the publication.

---

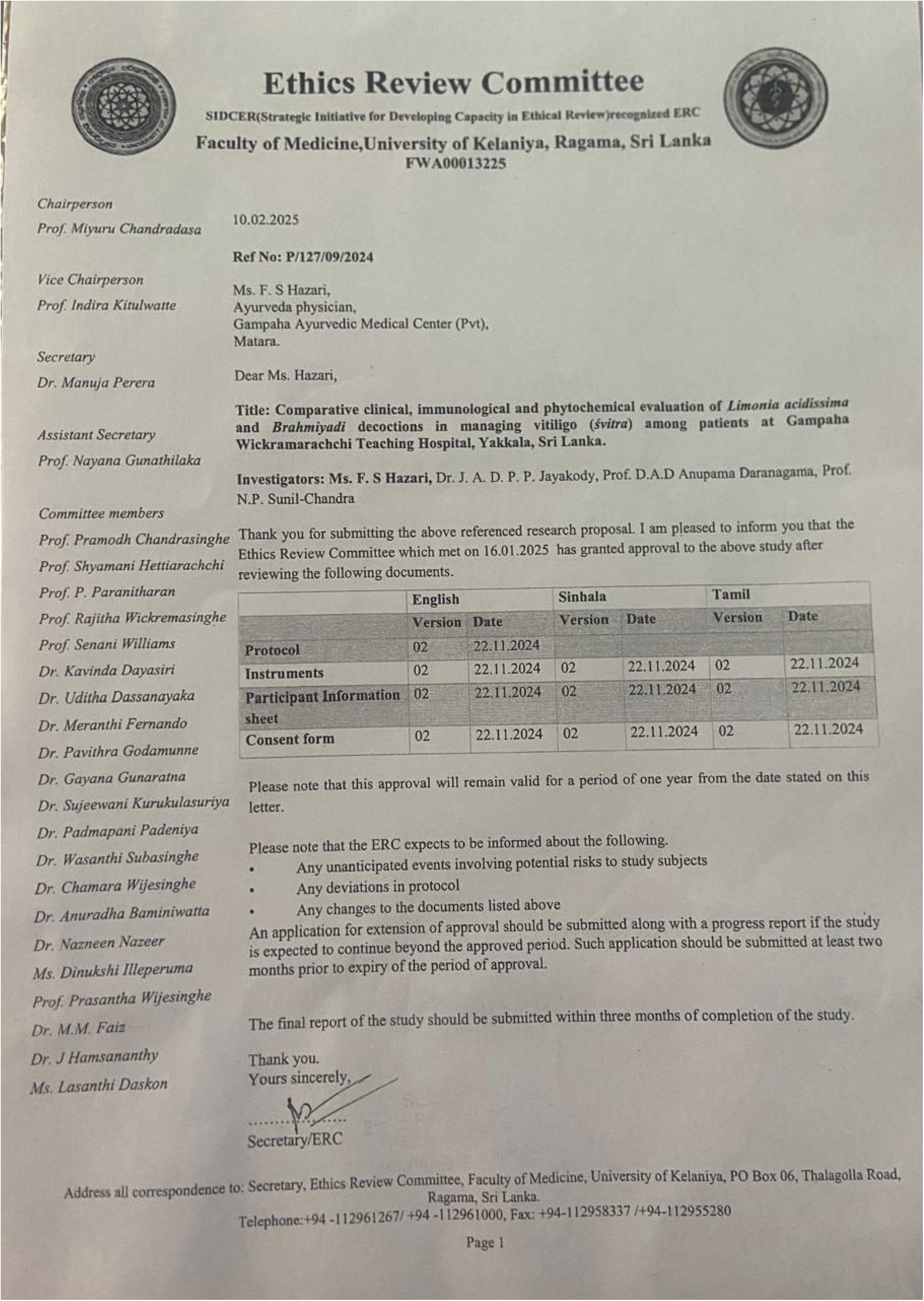

